# Risk and timing of miscarriage and stillbirth in five low- and middle-income countries: evidence from longitudinal cohort studies

**DOI:** 10.64898/2026.03.24.26349171

**Authors:** Zeleke Tadesse Joffe, Siaka Kone, Tefera Tesema, Irene Mugenya, Sailesh Mohan, Margaret E Kruk, Catherine Arsenault, Günther Fink, Emma Clarke-Deelder

**Affiliations:** Swiss Tropical and Public Health Institute, Allschwil, Switzerland; University of Basel, Basel, Switzerland; Centre Suisse de Recherches Scientifiques en Côte d’Ivoire, Abidjan, Côte d’Ivoire; Ethiopian Public Health Institute, Addis Ababa, Ethiopia; KEMRI-Wellcome Trust Research Program, Nairobi, Kenya; Centre for Chronic Disease Control, New Delhi, India; School of Medicine, Washington University in St. Louis, St. Louis, MO, USA; Department of Global Health, Milken Institute School of Public Health, The George Washington University, Washington, DC, USA

## Abstract

**Background:** Pregnancy loss, including miscarriage and stillbirth, is a major public health issue with major physical and psychological consequences for pregnant women. Prevalence estimates in low-resource settings remain scarce due to the lack of adequate data. This study assessed the prevalence, timing, and maternal characteristics associated with stillbirth and miscarriage using novel longitudinal data collected in five low- and middle-income countries (LMICs).

**Methods and Findings:** We analyzed longitudinal data from 5,755 pregnant women in Ethiopia, India, Kenya, South Africa, and Côte d’Ivoire. Women were enrolled during pregnancy and followed through delivery. Gestation-specific and cumulative risks of miscarriage and stillbirth were estimated using competing-risks survival analysis, adjusting for timing of enrollment. We examined associations with maternal age, education, wealth, and country using Fine and Gray sub-distribution hazard models. Among pregnancies surviving to 8 weeks, the cumulative risk of pregnancy loss by 28 weeks was 84 per 1,000 pregnancies (95% CI: 69–100) and from 28 to 44 weeks the risk was 19 per 1,000 (15–24), resulting in a total pregnancy loss risk after 8 weeks of gestation of 103 per 1,000 (88–119). Risks were highest in Côte d’Ivoire and lowest in South Africa. Losses peaked between 8 and 6 weeks of gestation, with a secondary rise after 36 weeks. Women aged ≥35 years had higher loss risk (HR 1.78, 95% CI: 1.27–2.48), whereas wealth and education showed no consistent association.

**Conclusions:** Pregnancy loss remains common across LMICs, with significant risk in both early and late gestation. Conventional estimates that do not account for delayed enrollment underestimate miscarriage rates. Enhanced surveillance and targeted interventions throughout pregnancy—especially during early gestation—are essential to reduce preventable fetal losses and meet associated global goals.

**Author’s Summary:** 

**Evidence before this study:**

- Data on the prevalence of miscarriage and stillbirth in low- and middle-income countries (LMICs) are scarce.
- Previous studies have reported substantial variation in miscarriage and stillbirth rates across LMICs, but most relied on retrospective surveys, facility-based data, or late pregnancy enrollment.
- Few multi-country longitudinal studies have examined gestation-specific patterns of pregnancy loss.

**Added value of this study:**

- This study uses newly available longitudinal cohort data from five LMICs and applies a novel competing risks survival methods to account for delayed enrollment and loss to follow-up.
- We show that failure to capture early pregnancy losses and failure to account for delayed entry and competing risksleads to systematic underestimation of miscarriage risk.
- We show that early pregnancy loss is substantially more common than suggested by crude estimates and that risk remains elevated beyond the first trimester.
- Our multi-country design allows for direct comparison across diverse health system and socioeconomic contexts.

**What do these findings mean?:**

- Our findings indicate that pregnancy loss in LMICs remains a major and under-estimated public health burden, particularly during early gestation.
- Improved measurement systems that capture pregnancies earlier and follow women longitudinally are essential for monitoring progress.
- Interventions to improve early antenatal care, strengthen risk screening, and ensure timely obstetric management are needed to reduce preventable losses.
- Policies focused solely on late pregnancy and delivery may miss a large proportion of fetal deaths occurring earlier in gestation.

## Background

Pregnancy loss, encompassing miscarriage and stillbirth, has profound physical, emotional, and economic consequences for affected women and their families. The risk of pregnancy loss is influenced by a range of factors including genetics, maternal health history, nutrition, and exposure to adversity during pregnancy (1–4). Pregnancy loss is often accompanied by pain, bleeding, and an urgent need for medical or surgical intervention, and may result in significant psychological trauma, increasing anxiety and stress during subsequent pregnancies (5). It is also associated with an elevated risk of common mental health disorders, such as depression and anxiety (6), and can negatively affect families’ economic well-being due to loss of income and heath care costs (5). In some cultural contexts, women who experience pregnancy loss may face social stigma or blame, further exacerbating psychological distress (7).

Despite the substantial burden of pregnancy loss, data on its prevalence are relatively limited. The gap is particularly pronounced in low- and middle-income countries (LMICs), where the burden of pregnancy loss is likely the highest but reliable data are often lacking (8). Until recently, national health surveys like the Demographic and Health Surveys did not capture information on the prevalence of miscarriages and stillbirths, and systematic pregnancies registries remain scarce. The World Health Organization (WHO) estimates that approximately 1.4% of pregnancies worldwide end in stillbirth after 28 weeks of gestation (9), but accurate estimation of stillbirth risk is hindered by limited vital registration systems, variation in stillbirth definitions, and gaps in the quality of routinely-collected health data (10). Measuring the risk of miscarriage is even more challenging because many pregnancies are lost before they are identified by the health system or included in research databases. A recent literature review estimated a pooled risk of miscarriage of 15.3% (95% CI: 12.5-18.7%) of all recognized pregnancies, but estimates from studies that enroll couples prior to conception find significantly higher rates ranging from 20% in a 2025 study in Canada and the United States (11) to 43% in a 1980 study in the United Kingdom (12). In high-income settings, chromosomal abnormalities account for the majority of early miscarriages; In lower income settings, the etiological profile may differ due to the higher burden of infectious diseases, maternal undernutrition, anemia, and limited access to early pregnancy care. Insufficient data on the prevalence and timing of pregnancy loss been identified as a critical gap, hindering progress toward global goals to reduce its burden (13).

In this study, we aimed to help address this critical gap by analyzing newly available data from the Maternal and Neonatal Health eCohort Study (MNH eCohort) in Ethiopia, Kenya, India, and South Africa (14), as well as longitudinal data from the Multigenerational Birth Cohort in Taabo, Côte d’Ivoire (Taabo MGC). The main objectives of this study were to estimate the timing and frequency of pregnancy loss across countries and assess variation in the risk of pregnancy loss by demographic and socioeconomic characteristics.

## Methods

### Study settings

This study analyzed data from pregnancy cohort studies in Côte d’Ivoire, Ethiopia, Kenya, India, and South Africa. Table 1 summarizes key demographic, economic, and health indicators for each country, highlighting the diversity of the populations studied. South Africa, an upper-middle-income country, has the highest Gross Domestic Product (GDP) per capita and the lowest maternal and neonatal mortality rates. In contrast, Ethiopia, a low-income country, has substantially higher mortality rates. India and Kenya, both lower-middle-income countries, show mixed profiles, with relatively strong maternal health service coverage alongside persistently high mortality burdens. Côte d’Ivoire, also classified as lower-middle-income, has higher neonatal, under-five, and maternal mortality rates than Kenya and India.

**Table 1:** Characteristics of five study countries.

| Indicator | Côte d’Ivoire | Ethiopia | India | Kenya | South Africa |
| --- | --- | --- | --- | --- | --- |
| GDP per capita (2023) | 2,531 | 1,272 | 2,481 | 1,952 | 6,023 |
| World Bank income group (2023) | Lower middle | Low | Lower middle | Lower middle | Upper middle |
| Population (Millions) (2025) | 33 | 135 | 1,464 | 58 | 65 |
| Neonatal mortality rate (per 1,000 live births) (2022) | 29 | 27 | 18 | 20 | 11 |
| Under-five mortality rate (per 1,000 live births) (2022) | 69.4 | 46.2 | 29.1 | 41.1 | 34.5 |
| Maternal mortality ratio (per 100,000 live births) (2023) | 359 | 195 | 80 | 149 | 118 |
| Rate of institutional delivery (2021) | 80.9 | 53.9 | 90.4 | 88.1 | 96.2 |

**Notes:** GDP stands for Gross Domestic Product. Estimates of GDP per capita, income group classifications, and estimates of maternal mortality ratios are from the World Bank Databank (15). Population estimates are from the Worldometer (16). Neonatal and under-5 mortality rates are from the World Health Organization (17). Institutional delivery rates are from the Demographic and Health Survey Programme (18).

### Inclusion criteria

Women were eligible for inclusion in the MNH eCohort if they were pregnant, aged 15 or above (18 or above in India), attending their first antenatal care visit at a selected study facility, planning to carry their pregnancy to term, and planning to reside in the study area for the duration of their pregnancy and follow-up. Women were eligible for inclusion in the Taabo MGC if they were pregnant and residing in the study area.

### Sampling and recruitment

In the MNH eCohort, women were sampled from the health facilities where they attended their first antenatal care visit. Facilities were categorized into four strata: public primary care facilities, public hospitals, private clinics, and private hospitals (where applicable). In India, only public primary and secondary care facilities were included, while in South Africa, the study was limited to public primary care facilities. Within strata, facilities were purposively selected with the goal of meeting recruitment targets within study timelines. Between 21 and 29 health facilities were selected in each country. The target sample size from each facility was determined with the goal of allocating the sample proportionally to different facility types based on past care-seeking patterns for antenatal care, using local health management information system data and Demographic and Health Surveys (DHS).

The MNH eCohort aimed to enroll a total of 1,000 pregnant women per country, with approximately 500 women enrolled from each of the two selected study sites. To achieve this target, a minimum of 50 women were recruited from each facility type, ensuring representation across public and private, as well as primary and hospital care settings. Data collectors worked alongside health providers to identify eligible women attending their first ANC visit at the study facilities.

The Taabo MGC aimed to enroll all eligible women living in the study area with an expected delivery date in 2024 or 2025 (a census of pregnant women). Women were recruited upon enrolment of their pregnancy into the routine pregnancy surveillance managed by the Health & Demographic Surveillance System (HDSS) in Taabo.

### Data sources and data collection

Across all cohorts, we analyzed data from baseline surveys and post-delivery follow-up surveys. In the MNH eCohorts, baseline surveys were conducted in-person with pregnant women at the time of their first antenatal care visit; in the Taabo MGC, baseline surveys were conducted in-person after enrolment and included information on their demographic characteristics and health history, and the estimated gestational age of their current pregnancy. Post-delivery follow-up surveys were conducted over the phone in the MNH eCohorts and in-person in the Taabo MGC. Follow-up surveys included information about when their pregnancy ended, and the outcome of the pregnancy.

Data collection for all cohorts was carried out using electronic data capture tools. Regular data audits were conducted to ensure the accuracy and completeness of the data, and any inconsistencies were addressed through follow-up verification with participants.

### Primary outcomes

The primary outcome for this study was pregnancy loss, including stillbirth and miscarriage. Miscarriage was defined as a fetal death before 28 weeks of gestation. Stillbirth was defined as a fetal death occurring at 28 weeks of gestation or later. In all cohorts, data on primary outcomes were collected during post-delivery follow-up interviews. To distinguish between stillbirth and neonatal death, interviewers used a standardized questionnaire to confirm the absence of breathing, crying, or movement at the time of delivery.

Gestational age was measured at the time of enrolment into each cohort study. In the MNH eCohort in Ethiopia, this was measured based on (1) the participant’s recall of the first day of her last menstrual period, or (2) if the participant did not remember the first day of her last menstrual period, her own report of her estimated gestational age, directly after her first antenatal care visit. In the MNH eCohorts in Kenya, India, and South Africa, this was measured based on the participant’s expected due date—self reported or extracted from her health record—at the time of enrolment on the day of her first antenatal care visit. Gestational age at enrolment was calculated as 40 minus the difference in weeks between the enrolment date and the expected due date. In the Taabo MGC, gestational age was calculated based on the first day of last menstrual period (LMP) and date of the end of pregnancy or based on the report on the stage of her pregnancy at the time of registration for respondents unable to provide reliable information on the date of LMP. To calculate gestational age at the end of pregnancy in the MNH eCohort, we calculated the time elapsed between enrolment and the end of pregnancy and added this time to the gestational age at enrolment. We used the same approach in the Taabo MGC, except for in the case of pregnancy loss; in these cases, we directly asked women about their gestational age at the of pregnancy loss.

In addition to the primary definitions aligned with WHO global reporting standards (miscarriage <28 weeks and stillbirth ≥28 weeks), we conducted secondary descriptive analyses using gestational age categories commonly used in clinical and high-income country reporting (early miscarriage <13 weeks, late miscarriage from 13 to <24 weeks, and stillbirth ≥24 weeks) to facilitate interpretation and comparison with other studies, as definitions of stillbirth vary internationally across thresholds ranging from approximately 20 to 28 weeks of gestation (19).

### Covariates

Maternal age was recorded as a continuous variable at the time of recruitment, based on participant self-report. We coded age into three categories: <20, 20-35, and over 35 years. Household wealth was assessed using a wealth index based on household assets and living conditions, using data collected at the time of enrolment. In the MNH eCohort countries, this included: ownership or a radio, television, telephone, refrigerator, car, motorbike, bicycle; having a bank account, having access to a safe water source, having an improved toilet, flooring material, wall material, and roofing material. In the Taabo MGC, this included air conditioning, bike, car, computer, electricity, fan, fridge internet, iron, mobile phone, motorbike, TV, water pump. This index was calculated separately in each study country and categorized into within-sample tertiles, with higher categories representing greater wealth.

We coded the reported education levels into three categories: none or some primary, completed primary, or completed secondary education. We coded location into two categories: urban or rural. In each of the three African countries, two sentinel sites were selected—one predominantly rural and one urban. In India, the study was conducted in two districts from two different states: Sonipat district in Haryana and Jodhpur in Rajasthan; facilities were classified as being in urban or rural areas within these two districts. In the Taabo MGC, participants were categorized according to whether they lived in the main urban area of Taabo (Taabo-cité) or in the surrounding rural areas.

### Statistical analysis

We began our analysis by describing the characteristics of women in the study samples from each of the five participating countries and in the pooled sample. We created two samples for analysis. First, for descriptive analyses, we restricted our sample to women with information on gestational age at the time of enrolment and with complete follow-up through the end of pregnancy (“complete follow-up sample”). Second, for survival analysis that accounts for loss to follow-up, we defined a sample that includes all women with information on gestational age at the time of enrolment and at least one follow-up interview during pregnancy (“full analytic sample”). We excluded women with induced abortions from both samples.

We calculated the proportion of women in the complete follow-up sample who experienced miscarriage or stillbirth. However, these crude proportions are biased estimates of the true risk. This bias arises for two reasons. First, the complete follow-up sample excludes women who were lost to follow-up, a group that may differ systematically in their risk of pregnancy loss. Second, participants enrolled at different gestational ages, and those who enrolled later – often not until the second trimester—contributed no person-time at risk during early gestation. As a result, crude estimates disproportionately reflect later gestation and systematically underrepresent early pregnancy losses, leading to an underestimation of the overall risk.

To address the issues of varying enrolment timing and loss-to-follow-up, we estimated the risk of pregnancy loss by gestational age and overall using survival analysis techniques. We focused on pregnancy loss conditional on survival to 8 weeks of gestation, due to sparse data prior to eight weeks.

To estimate cumulative risks of pregnancy loss over gestation, we conducted a competing risks analysis and constructed cumulative incidence curves. Live birth precludes the subsequent occurrence of pregnancy loss and therefore represents a competing event rather than a censoring event. Treating live birth as a competing risk, rather than censoring at birth, avoids the upward bias in cumulative risk estimates that would result from standard survival methods that assume non-informative censoring (20). In this analysis, observations were censored at the date of last contact or at 44 weeks after the last menstrual period. We reported results through 44 weeks’ gestation because we observed a nontrivial number of pregnancy losses through this point; this may be partly driven by error estimation in gestational age, as pregnancy through 44 weeks’ gestation is unlikely.

Using cumulative incidence functions (CIFs), we estimated cumulative risks of pregnancy loss, neonatal death, and their combination for each country and for the pooled sample, treating live birth as a competing risk. We used CIFs to estimate the risk of miscarriage, defined as the cumulative risk of pregnancy loss by 28 weeks gestation, and the risk of stillbirth, defined as the cumulative risk from 28 weeks’ gestation to 44 weeks’ gestation. Confidence intervals for cumulative incidence at a fixed gestational age (e.g., miscarriage risk) were obtained directly from the CIF, whereas confidence intervals for differences between two points on the CIF (e.g., stillbirth risk) were estimated using a nonparametric bootstrap with re-estimation of the CIF in each resampled dataset. In addition, we estimated stillbirth risk as a proportion of births, excluding miscarriages from the denominator to align with international definitions of stillbirth.

We additional estimated gestational-age-specific prospective risks of pregnancy loss using “fetuses-at-risk” perspective (21). We calculated the risk of pregnancy loss in four-week time intervals of gestational age, among fetuses at risk at the beginning of the time interval. We did this using the following formula:

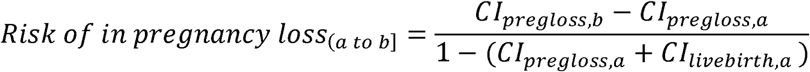

where *CI_pregloss,x_* and *CI_livebirth,x_* represent the cumulative incidence of pregnancy loss and live birth, respectively, up to gestational age *x*. These estimates provide information about the risk of pregnancy loss in a given time interval among fetuses who have survived to the start of this interval.

Finally, we examined variation in pregnancy loss by maternal characteristics, using a Fine and Gray sub-distribution hazard model for competing risks (22). Models were adjusted for maternal age, education, and wealth tertile. We first estimated models individually by country, and then estimated a pooled model adjusted for country fixed effects in addition to individual covariates.

#### Ethics

Ethical approval for the original data collection was obtained in all countries (23,24). This analysis used only de-identified or anonymized secondary data provided to the authors.

#### Data availability

Data will be made available by the authors upon request. The analysis code will be published with the paper (supplemental materials).

#### Funding

No funding was obtained for this project. The E-cohorts study was funded through a grant to the QuEST network by the Gates Foundation (grant number INV-005254) as well as the Swiss Development Corporation (grant number 81067262). The Taabo MGC project was funded through the Eckenstein-Geigy Professorship at the University of Basel.

## Findings

Data were collected from 6057 participants, including 1980 participants from Cote d’Ivoire, 1000 from Ethiopia, 1023 participants from India, 1009 from Kenya, and 1045 from South Africa. The final analytic sample included 5755 participants. Some participants were lost to follow-up before the end of their pregnancies or had missing information on gestational age at the time of pregnancy outcome. Complete follow-up data was available for 5463 participants. **Figure S1** shows the sample flow chart.

India had a predominantly urban participant sample, with approximately 64.3% of participants residing in urban areas. In contrast, Kenya, Ethiopia, and South Africa had a more balanced rural-urban distribution, with nearly equal proportions of participants from both settings. Côte d’Ivoire had a predominantly rural sample, with 75.3% of participants residing in rural areas. Regarding the timing of enrollment, most participants in sample in Ethiopia, Kenya, South Africa and Côte d’Ivoire were enrolled during the second trimester of pregnancy. India had the highest proportion of participants enrolling in the first trimester (53.0% of the complete follow-up sample) (**Table 2**).

**Table 2:** Participant characteristics.

|  | **Côte d'Ivoire** | **Ethiopia** | **India** | **Kenya** | **South Africa** | **Pooled** |
| --- | --- | --- | --- | --- | --- | --- |
|  | **N=1922** | **N=864** | **N=956** | **N=859** | **N=861** | **N=5462** |
|  | N (%) | N (%) | N (%) | N (%) | N (%) | N (%) |
| **Site** |  |  |  |  |  |  |
| Urban | 473 (24.6) | 430 (49.8) | 603 (63.1) | 401 (46.7) | 425 (49.4) | 2345 (42.6) |
| Rural | 1449 (75.4) | 434 (50.2) | 353 (36.9) | 458 (53.3) | 436 (50.6) | 3159 (57.4) |
| **Trimester at enrollment** |  |  |  |  |  |  |
| 1st trimester | 79 (4.1) | 263 (30.4) | 507 (53.0) | 146 (17.0) | 340 (39.5) | 1337 (24.3) |
| 2nd trimester | 1236 (64.3) | 554 (64.1) | 397 (41.5) | 561 (65.3) | 430 (49.9) | 3203 (58.2) |
| 3rd trimester | 607 (31.6) | 47 (5.4) | 52 (5.4) | 152 (17.7) | 91 (10.6) | 963 (17.5) |
| **Highest level of education** |  |  |  |  |  |  |
| None or some primary | 677 (35.2) | 319 (36.9) | 146 (15.3) | 65 (7.6) | 7 (0.8) | 1937 (35.2) |
| Completed primary | 652 (33.9) | 351 (40.6) | 311 (32.5) | 296 (34.5) | 250 (29.0) | 1712 (31.1) |
| Completed secondary or higher | 596 (31.0) | 194 (22.5) | 500 (52.3) | 498 (58.0) | 604 (70.2) | 1855 (33.7) |
| **Age at enrollment** |  |  |  |  |  |  |
| Age <20 | 344 (17.9) | 69 (8.0) | 69 (7.2) | 101 (11.7) | 139 (16.1) | 721 (13.1) |
| Age 20-34 | 1226 (63.8) | 739 (85.5) | 868 (90.8) | 633 (73.7) | 598 (69.4) | 4095 (74.4) |
| Age 35+ | 352 (18.3) | 56 (6.5) | 19 (2.0) | 125 (14.6) | 126 (14.6) | 682 (12.4) |
| **Pregnancy characteristics** |  |  |  |  |  |  |
| Multiple gestation | 42 (2.2) | 16 (2.0) | 6 (0.6) | 18 (2.1) | 19 (2.2) | 101 (1.8) |

Notes: Table shows the characteristics of participants with complete follow-up. Sample statistics are shown at the level of the pregnant woman.

In the samples from South Africa, India, and Kenya, most women had completed secondary education. In contrast, only a minority of women in Ethiopia (22.5%) and Côte d’Ivoire (2.9%) had secondary education. In all five study countries, most participants were aged between 20 and 34 years. The proportion of adolescent participants (<20 years) ranged from 7.2% in India to 17.7% in Côte d’Ivoire.

In the complete follow-up sample, without adjusting for differential enrolment timing, 177 (3.2%) women experienced a miscarriage and 114 (2.1%) experienced a stillbirth. The percentage experiencing a miscarriage ranged from 2.0% in Ethiopia to 5.1% in South Africa, and the percentage experiencing a stillbirth ranged from 1.3% in South Africa to 3.0% in Côte d’Ivoire (**Table S2**).

**Figure 1** presents the cumulative incidence of pregnancy loss by gestational age across the five study countries. The highest risk of pregnancy loss occurs during the earliest weeks of gestation, with the steepest increases in mortality observed between 8 and 16 weeks. The survival curves flatten during mid-gestation (16–40 weeks), and then increases again towards the end of gestation, particularly beyond 40 weeks. By the end of gestation, cumulative risk is by far highest in Cote d’Ivoire, and lowest in the sample from South Africa.

**Figure 1:**
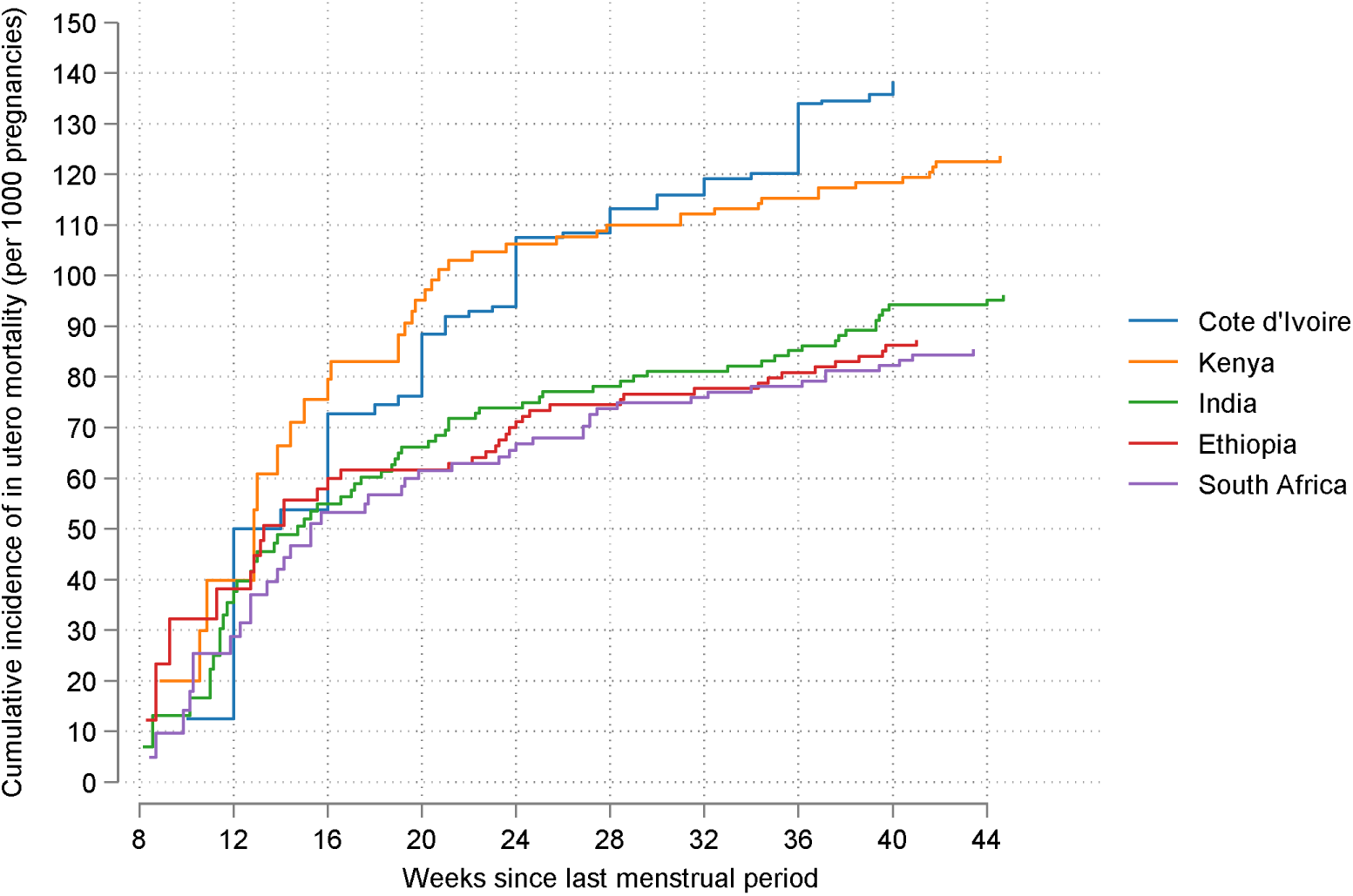
Cumulative incidence of pregnancy loss by country.

**Figure 2** shows the cumulative incidence of pregnancy loss in relation to other birth outcomes (live birth, ex utero mortality, and all fetal mortality and ex utero mortality) in the pooled 5-country sample. This figure demonstrates the relative amount of in-utero to ex-utero mortality through 44 weeks of gestational age. In the pooled sample, between 36 weeks and 44 weeks of gestational age, 80% of mortality occurs in utero (as stillbirths) and 20% occurs ex utero (as newborn or infant deaths).

**Figure 2:**
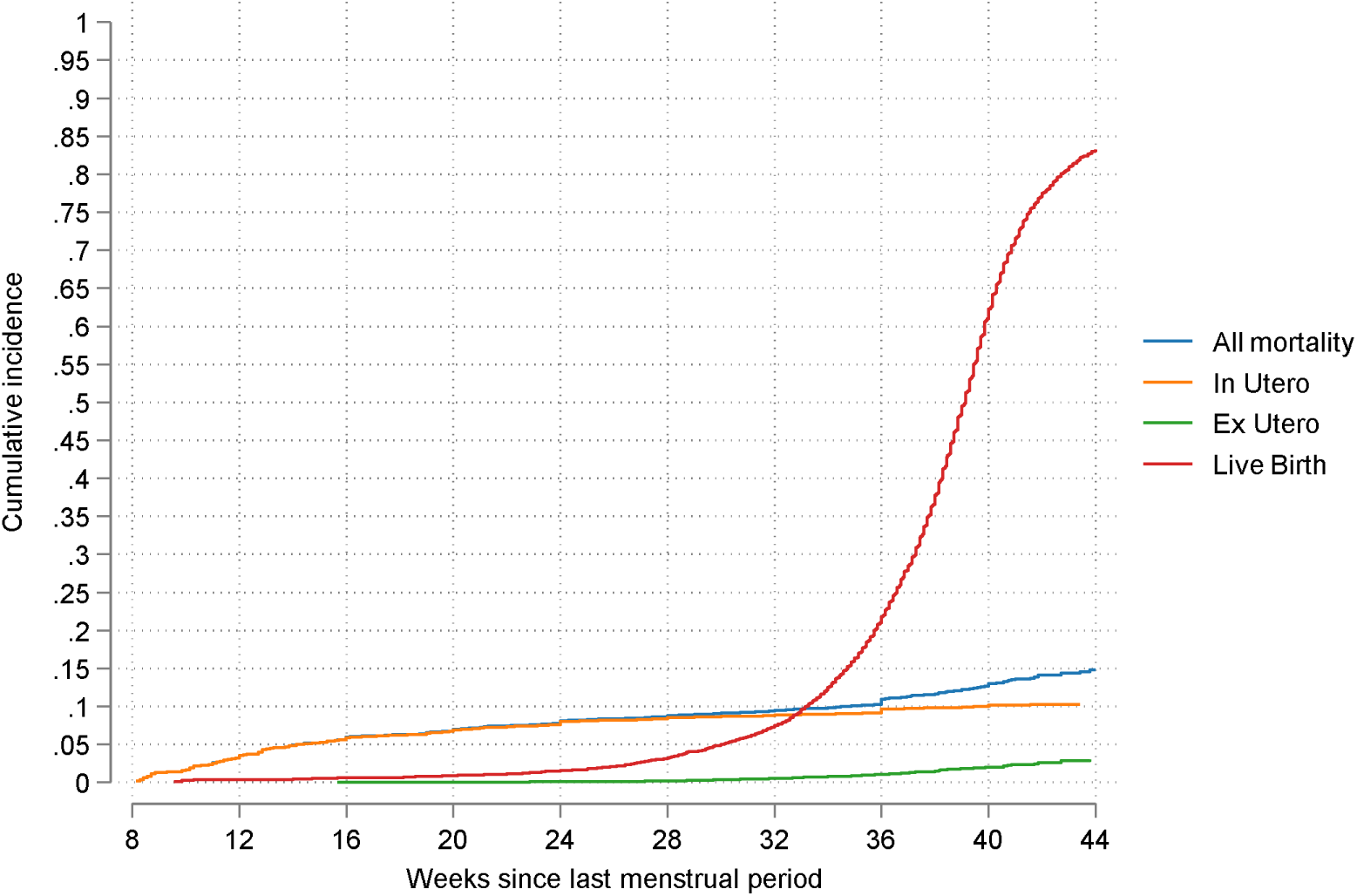
Cumulative incidence of pregnancy loss, live birth, neonatal mortality, and all mortality in the pooled 5-country sample.

**Table S4** shows gestational age-specific pregnancy loss rates using a fetuses-at-risk approach. Across all samples, the highest risk period were early in pregnancy. Among fetuses-at-risk at 8 weeks of gestation, the risk of mortality from 8-12 weeks ranged from 29 fetal deaths per 1000 fetuses-at-risk in India to 50 fetal deaths per 1000 fetuses at risk in Cote d’Ivoire. From 12 to 16 weeks’ gestation, the risk ranged from 21 fetal deaths per 1000 fetuses at risk in South Africa and Ethiopia to 38 fetal deaths per 1000 fetuses at risk in Kenya. The risk decreased over the course of the second trimester and beginning of the third trimester and then, in all countries, increased after 32 weeks of gestation.

**Table 3** presents the cumulative risk of pregnancy loss by 28 weeks, from 28 weeks to 44 weeks, and by 44 weeks gestation among pregnancies that survive to 8 weeks, in each study country and overall, calculated based on the competing risks analysis. By 28 weeks of gestation, the cumulative risk ranged from 74 per 1000 pregnancies in Ethiopia and South Africa, to 108 per 1000 pregnancies in Côte d’Ivoire, with a pooled estimate of 84 per 1000 pregnancies (95% CI: 69,100). From 28 weeks to 44 weeks, the risk was lowest in South Africa at 12 deaths per 1000 pregnancies (95% CI: 3, 21) and highest in Côte d’Ivoire at 30 deaths per 1000 pregnancies (95% CI: 20, 40). Through 44 weeks, the cumulative risk of pregnancy loss reached 138 per 1000 pregnancies (95% CI: 94, 191) in Côte d’Ivoire, and 103 per 1000 pregnancies (95% CI: 88, 119) in the pooled analysis. These findings highlight both inter-country variability and the increasing burden of pregnancy loss as gestation progresses. Côte d’Ivoire and Kenya consistently showed higher risks, suggesting potential contextual factors influencing fetal outcomes that warrant further investigation.

**Table 3:** Fetal and neonatal mortality rates by country.

| Country | Pregnancy loss up to 28 weeks gestation per 1,000 pregnancies | Pregnancy loss between 28 weeks and 44 weeks gestation per 1,000 pregnancies | Pregnancy loss up to 44 weeks gestation per 1,000 pregnancies | Neonatal mortality per 1,000 live births |
| --- | --- | --- | --- | --- |
| Ethiopia | 74 (41, 121) | 13 (3, 22) | 87 (53, 133) | 16 (8, 25) |
| Kenya | 110 (63, 171) | 13 (3, 22) | 123 (75, 182) | 19 (10, 28) |
| South Africa | 74 (50, 104) | 12 (3, 21) | 85 (61, 116) | 8 (2, 14) |
| India | 78 (54, 108) | 17 (6, 28) | 95 (70, 125) | 13 (6, 21) |
| Côte d'Ivoire | 108 (65, 164) | 30 (20, 40) | 138 (94, 191) | 22 (16, 29) |
| Pooled | 84 (69, 100) | 19 (15, 24) | 103 (88, 119) | 17 (14, 21) |

**Note:** Pregnancy loss risks are calculated from cumulative incidence curves estimated through a competing risks model. Risks of pregnancy loss are presented per 1,000 identified pregnancies, conditional on survival to 8 weeks of gestation. Neonatal mortality is the number of deaths within 28 days per 1,000 live births. 95% confidence intervals are shown in parentheses.

Using alternative definitions, in the pooled sample, we estimated risks of loss from 8 to <13 weeks of 43 (31, 59), from 13 to <24 weeks of 33 (23, 43) and from 24 weeks onwards of 27 (31, 32) per 1000 pregnancies (**Table S3**).

**Table 4** shows the result from competing risks regression models analyzing factors associated with pregnancy loss. Women aged 35 years or older had a 78 percent higher hazard of pregnancy loss (95% CI: 1.27, 2.48, p=0.001). We found no association with wealth or maternal education. Relative to the women in the Ethiopian sample, women in Cote d’Ivoire had a substantially increased pregnancy loss risk (HR 1.94, [1.30, 2.89], p<0.01).

**Table 4:**
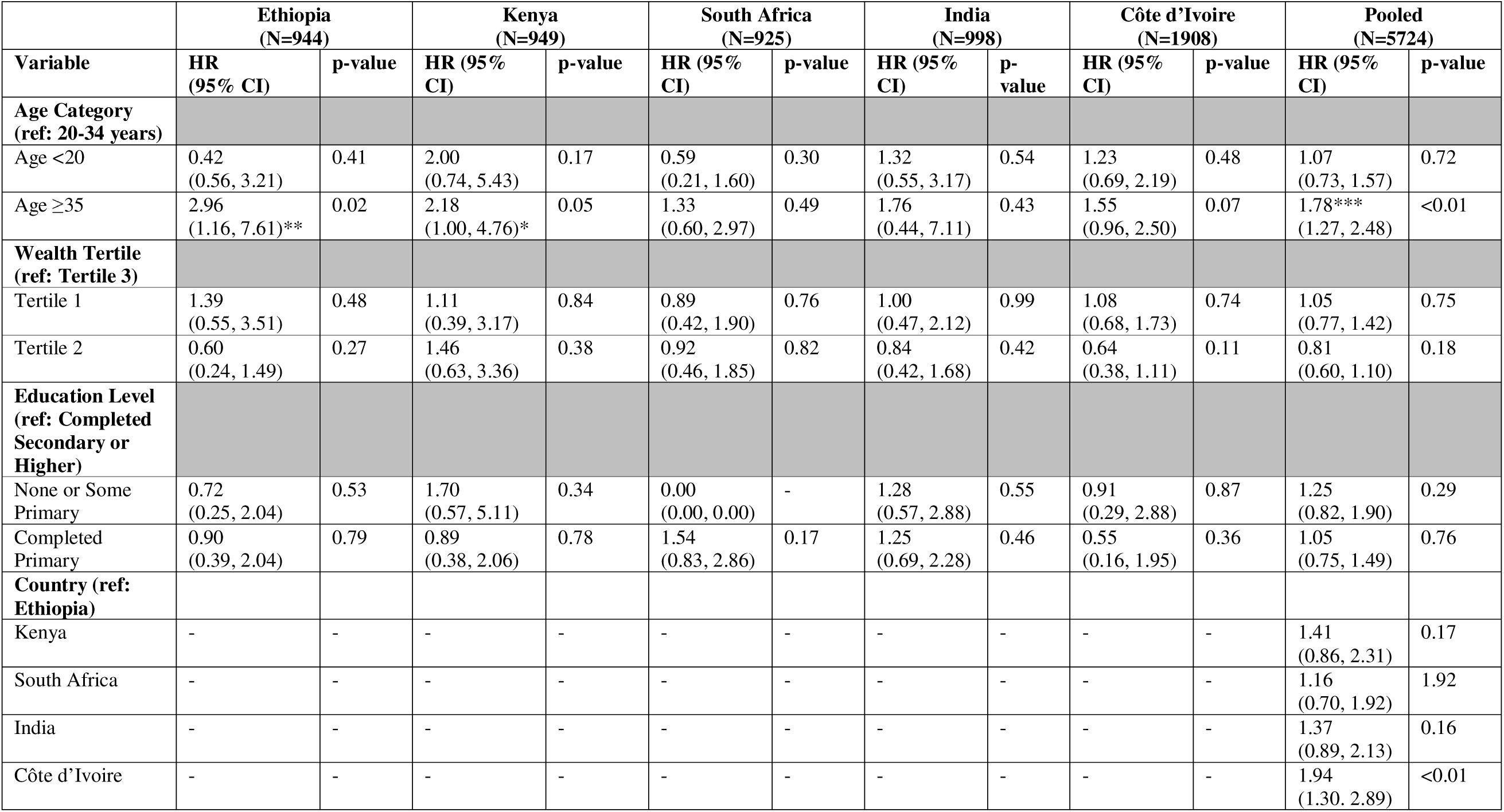
Competing risks regression for pregnancy loss in five low- and middle-income countries.

## Discussion

Using a novel data set covering 5755 pregnancies from Côte d’Ivoire, Ethiopia, India, Kenya, and South Africa we show that on average 103 out of 1000 pregnancies are lost between gestational weeks 8 and 44, with 84 per 1000 pregnancies lost between gestational weeks 8 and 28, and 19 lost at gestational week 28 or later. The risk of pregnancy loss appears highest among women over age 35, and we observe significant variation across countries.

Our estimates of miscarriage risk are generally higher than those in published literature. We conducted a meta-analysis of past studies of the risk of miscarriage across Cote d’Ivoire (25), Ethiopia (26,27), India (28,29), Kenya (30,31), and South Africa (32,33) (**Figure S2**) and estimated a pooled miscarriage risk of 56 per 1000 pregnancies. By contrast, our estimate is 84 per 1000 pregnancies. We believe this difference is primarily driven by methodological approaches to handling late enrollment. Many studies estimate miscarriage risk by calculating the fraction of participants who miscarried after enrolment, not accounting for the selection that occurs when women enroll late in pregnancy. If we just look at survival among enrolled women, we estimate a risk of 35 miscarriages per 1000 pregnancies, which is less than half of our preferred estimate. While it is impossible to capture all pregnancies from the very beginning, accounting for early selection appears critical for obtaining consistent and reliable data.

When we calculate the stillbirth rate with miscarriages removed from the denominator, to align with the internationally-used stillbirths definition of deaths per 1000 live births, we estimate rates of: 14 stillbirths per 1000 live births in Ethiopia, 15 per 1000 in Kenya, 13 per 1000 in South Africa, 18 per 1000 in India, and 34 per 1000 live births in Côte d’Ivoire. The pooled rate is 21 stillbirths per 1000 live births. Our estimates of stillbirth risks are very similar to those found in the published literature from the study countries (25–33). The pooled estimate from our meta-analysis of past studies in the five study countries was 23 per 1000 births (**Figure S3**). Estimates of the effect of stillbirth are less influenced by enrolment timing, given that stillbirths occur late in pregnancy.

Stillbirth rates observed in our study remain substantially higher than those typically reported in high-income countries. For example, a review of stillbirth definitions and reporting practices in Europe reported a median stillbirth rate of approximately 2.7 per 1000 births when using the ≥28-week definition, increasing to 3.3 per 1000 births when deaths occurring between 24 and 27 weeks were included (34). These substantially lower rates in high-income settings likely reflect stronger health systems, including broader access to antenatal care, improved detection of maternal and fetal complications, and higher quality intrapartum care.

Pregnancy loss rates varied considerably across countries. The overall pregnancy loss risk was highest in Côte d’Ivoire and lowest in South Africa while the risk of miscarriage was highest in Kenya and Côte d’Ivoire, and lowest in South Africa. The risk of stillbirth was highest in Côte and lowest in Kenya and South Africa. This variation may be attributed to differences in underlying risk factors, health behaviors, and healthcare access that likely depend largely on local context. We find a surprisingly weak overall relationship between maternal characteristics and these outcomes.

Across the study countries, the risk of pregnancy loss was highest from 8-12 weeks, with a marked decline after 16 weeks of gestation. Fetus-at-risk analyses and survival curves revealed that conditional on survival to 8 weeks of gestation, the highest loss rates occurred between 8–16 weeks, with early pregnancy losses being more common than late miscarriages or stillbirths. Our findings align with previous research indicating that early pregnancy loss is more common than losses occurring in later stages of pregnancy, particularly up to 12 weeks of gestation (42, 48). However, our study extends this understanding by revealing that pregnancy loss remains notably high beyond 12 weeks and continues to be substantial up to 16 weeks of gestation.

A secondary peak in fetal loss beyond 36 weeks was observed in some settings, suggesting a potential contribution of post-term complications, inadequate intrapartum care, and undiagnosed maternal-fetal complications. These findings highlight the importance of early prenatal care and timely delivery interventions to prevent adverse outcomes.

One of the strengths of this study is its reliance on longitudinal data from multiple LMICs, providing a robust comparison of pregnancy loss trends across diverse settings. The use of survival analysis techniques allowed for a nuanced understanding of gestation-specific risks and the impact of maternal characteristics.

However, some limitations must be acknowledged. First, gestational age estimates often relied on maternal recall of the last menstrual period (LMP), which may be prone to misclassification bias, particularly in settings with limited access to early ultrasound. This could have affected the classification of miscarriage versus stillbirth and the estimated timing of pregnancy losses. However, estimates of the overall risk of pregnancy loss are not affected by measurement of gestational age.

Second, we report estimates conditional on survival to 8 weeks’ gestational age due to sparse data from earlier in pregnancy. As a result, we omit the highest-risk period of pregnancy. Studies that include this early high-risk period – which requires enrolment prior to conception and frequent pregnancy testing of all participants – remain scarce but report much higher risks of pregnancy loss, in the range of 330 to 430 losses per 1000 pregnancies (26, 27).

Third, we do not know the pregnancy outcomes of participants who were lost to follow-up before the end of their pregnancies or did not have information on gestational age at the end of pregnancy. Our analysis assumes that these participants faced the same risk across pregnancy weeks as participants with complete follow-up. This assumption may not hold if loss to follow-up is associated with underlying characteristics that are associated with pregnancy loss risk. However, our study had a relatively low rate of loss-to-follow-up, ranging from 3% in Cote d’Ivoire to 19% in South Africa.

## Conclusion

Our findings highlight a major gap in progress toward the global goal of ending preventable newborn deaths and stillbirths, as outlined in Every Newborn: An Action Plan to End Preventable Deaths by 2035. The persistently high burden of pregnancy loss in LMICs underscores the urgent need for increased investment in maternal and newborn health programs, particularly in strengthening antenatal care, improving access to quality obstetric services, and addressing underlying risk factors. Without targeted interventions and policy shifts, the current trajectory suggests that many countries will struggle to meet the ambitious targets set for 2035.

## Supplementary Appendix

**Figure S1:**
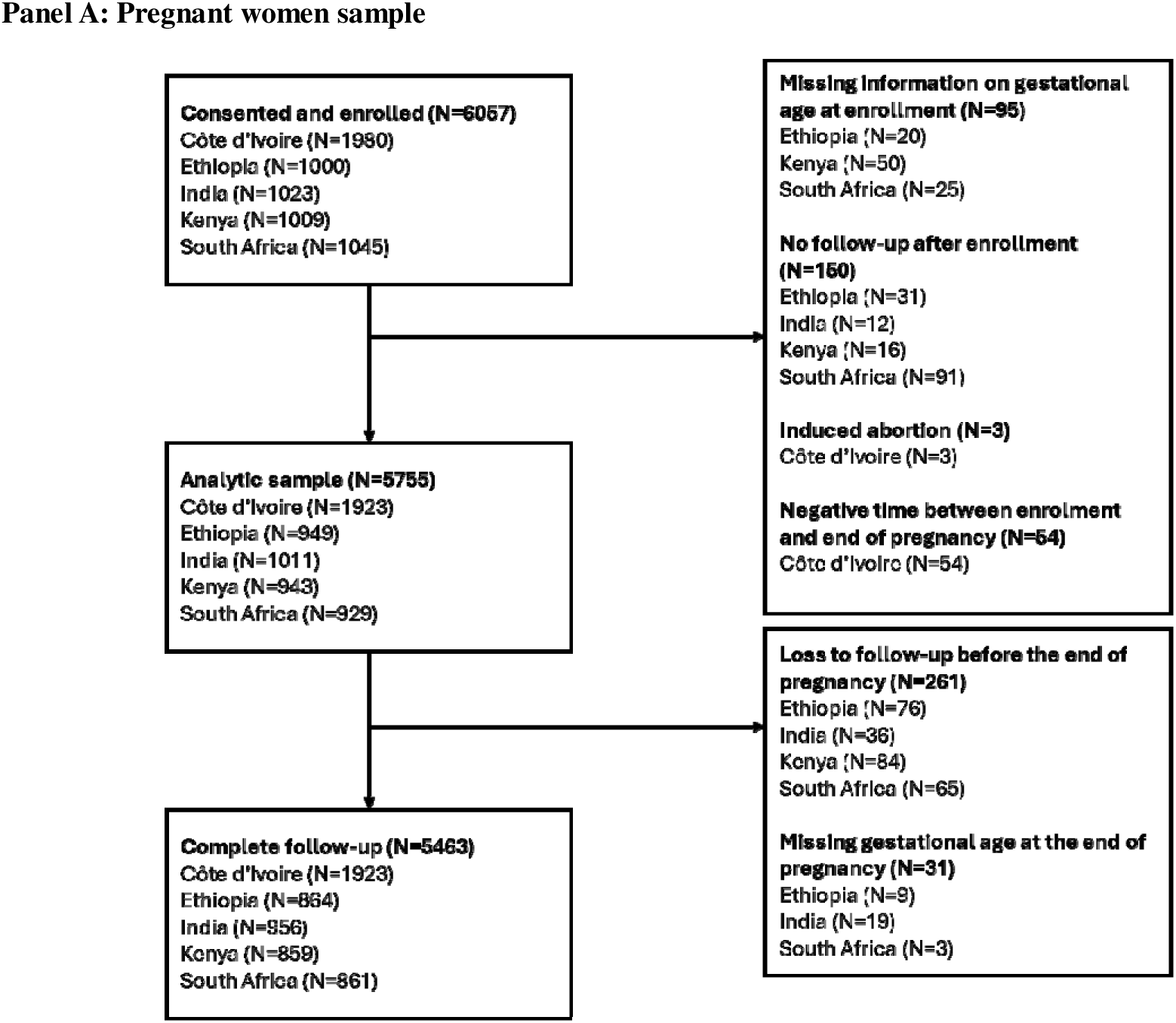
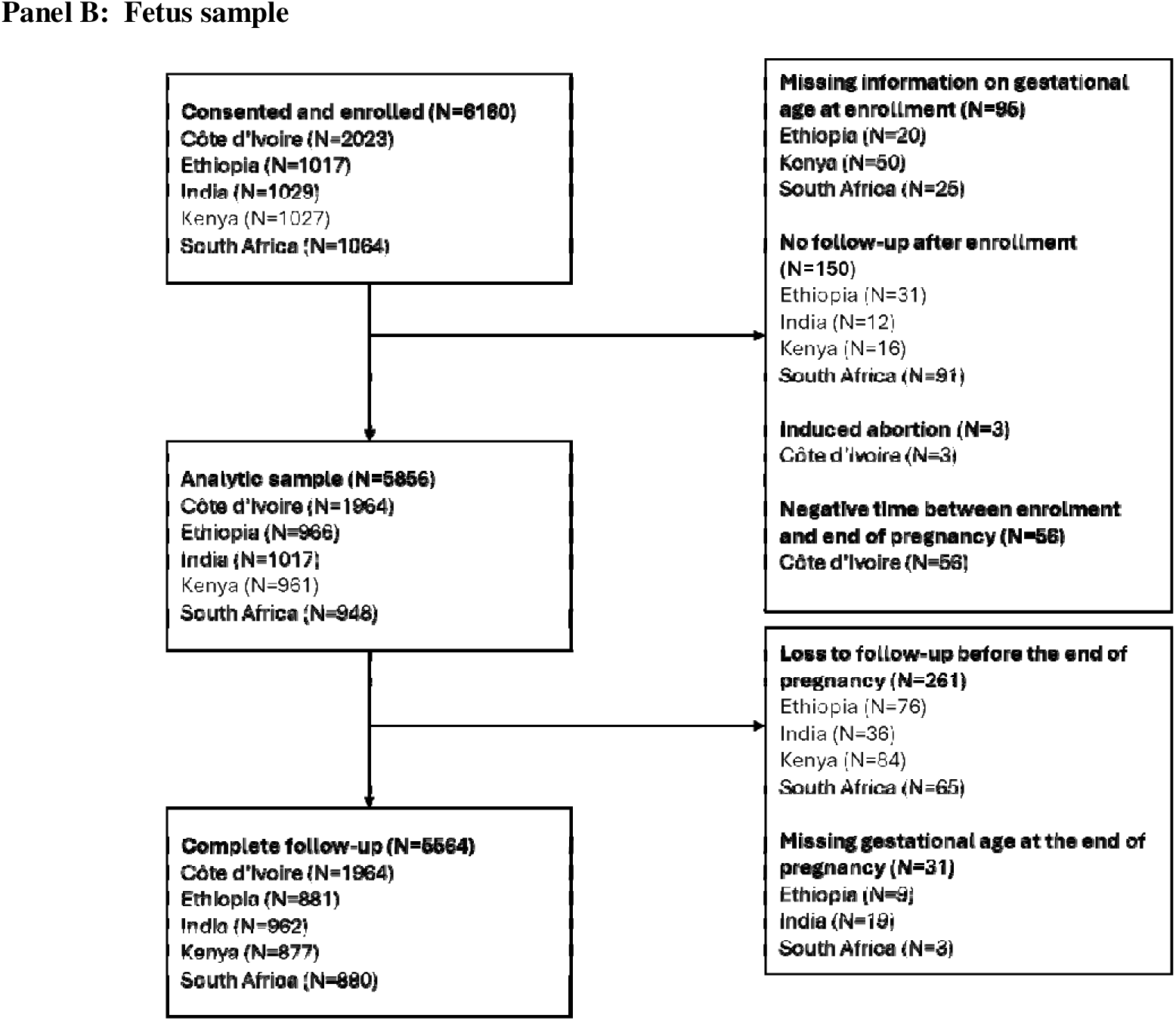
Study sample diagram.

**Table S1:**
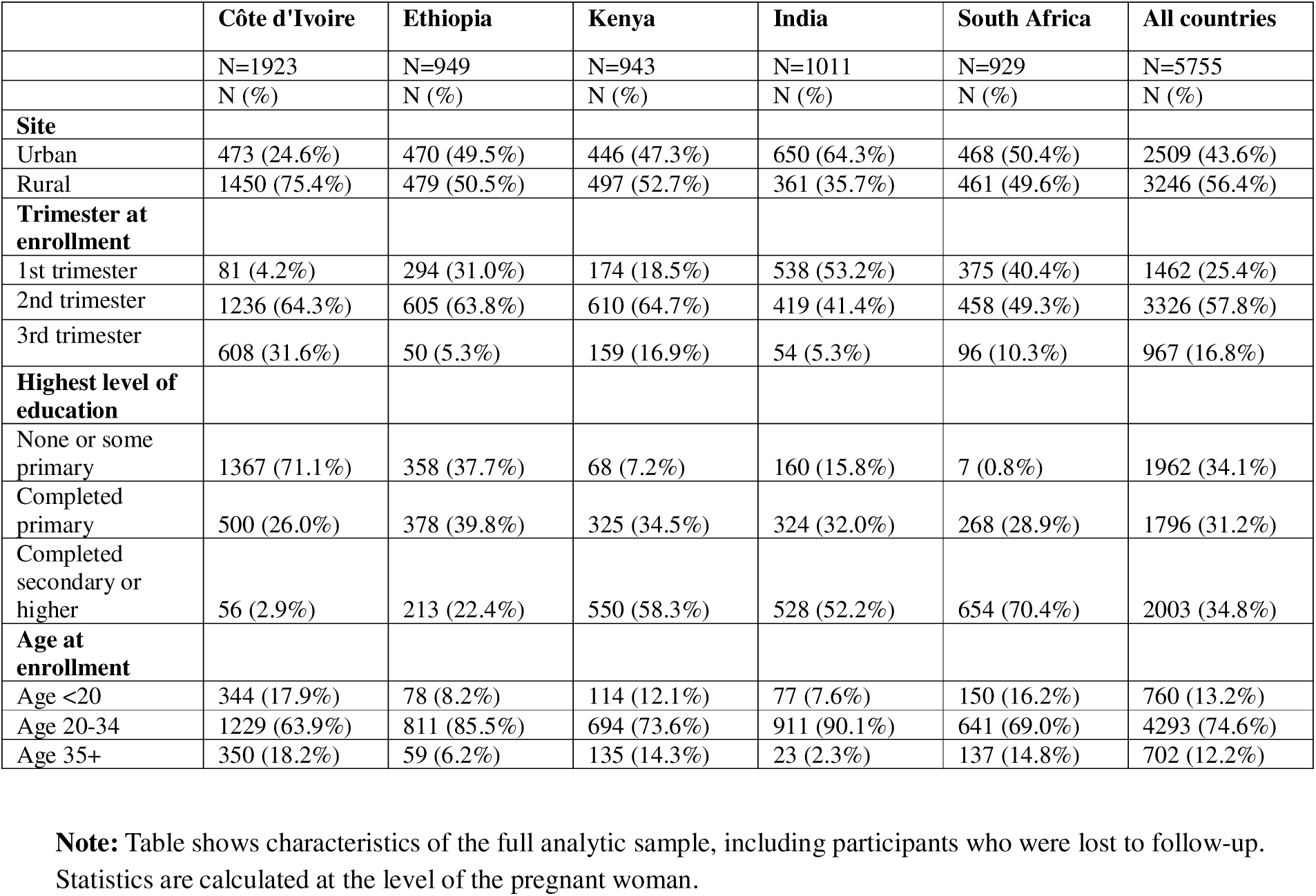
Participant characteristics for the full analytic sample.

**Table S2:**
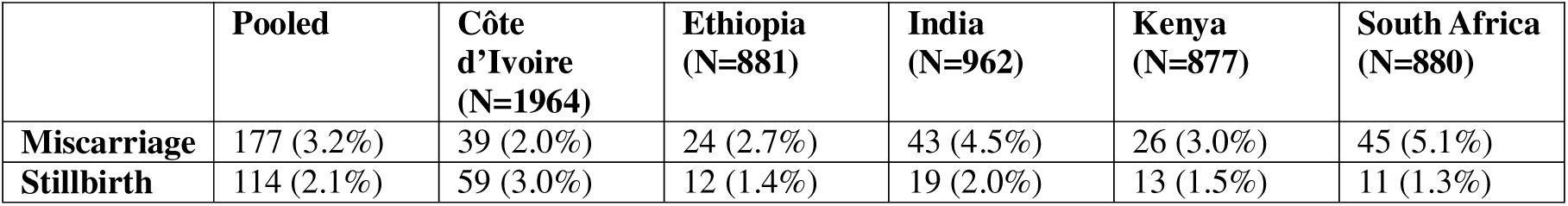
Pregnancy outcomes among women followed-up through the end of pregnancy, not adjusted for left truncation.

**Table S3:**
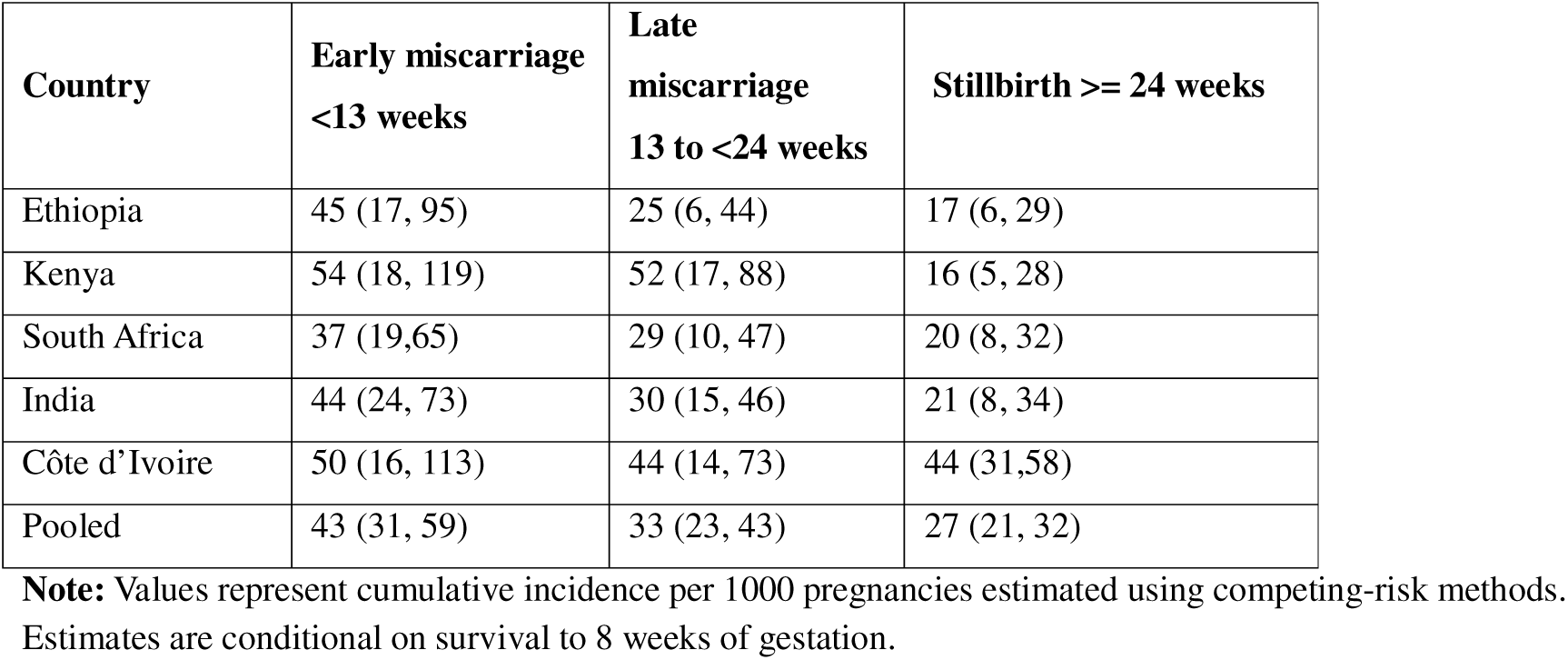
Cumulative incidence of pregnancy loss at selected gestational ages, conditional on survival to 8 weeks of gestation, by country (alternative definitions)

| **Country** | **Early miscarriage**<br>**<13 weeks** | **Late**<br>**miscarriage**<br>**13 to <24 weeks** | **Stillbirth ≥ 24 weeks** |
| --- | --- | --- | --- |
| Ethiopia | 45 (17, 95) | 25 (6, 44) | 17 (6, 29) |
| Kenya | 54 (18, 119) | 52 (17, 88) | 16 (5, 28) |
| South Africa | 37 (19,65) | 29 (10, 47) | 20 (8, 32) |
| India | 44 (24, 73) | 30 (15, 46) | 21 (8, 34) |
| Côte d'Ivoire | 50 (16, 113) | 44 (14, 73) | 44 (31,58) |
| Pooled | 43 (31, 59) | 33 (23, 43) | 27 (21, 32) |

**Note:** Values represent cumulative incidence per 1000 pregnancies estimated using competing-risk methods. Estimates are conditional on survival to 8 weeks of gestation.

**Figure S2:**
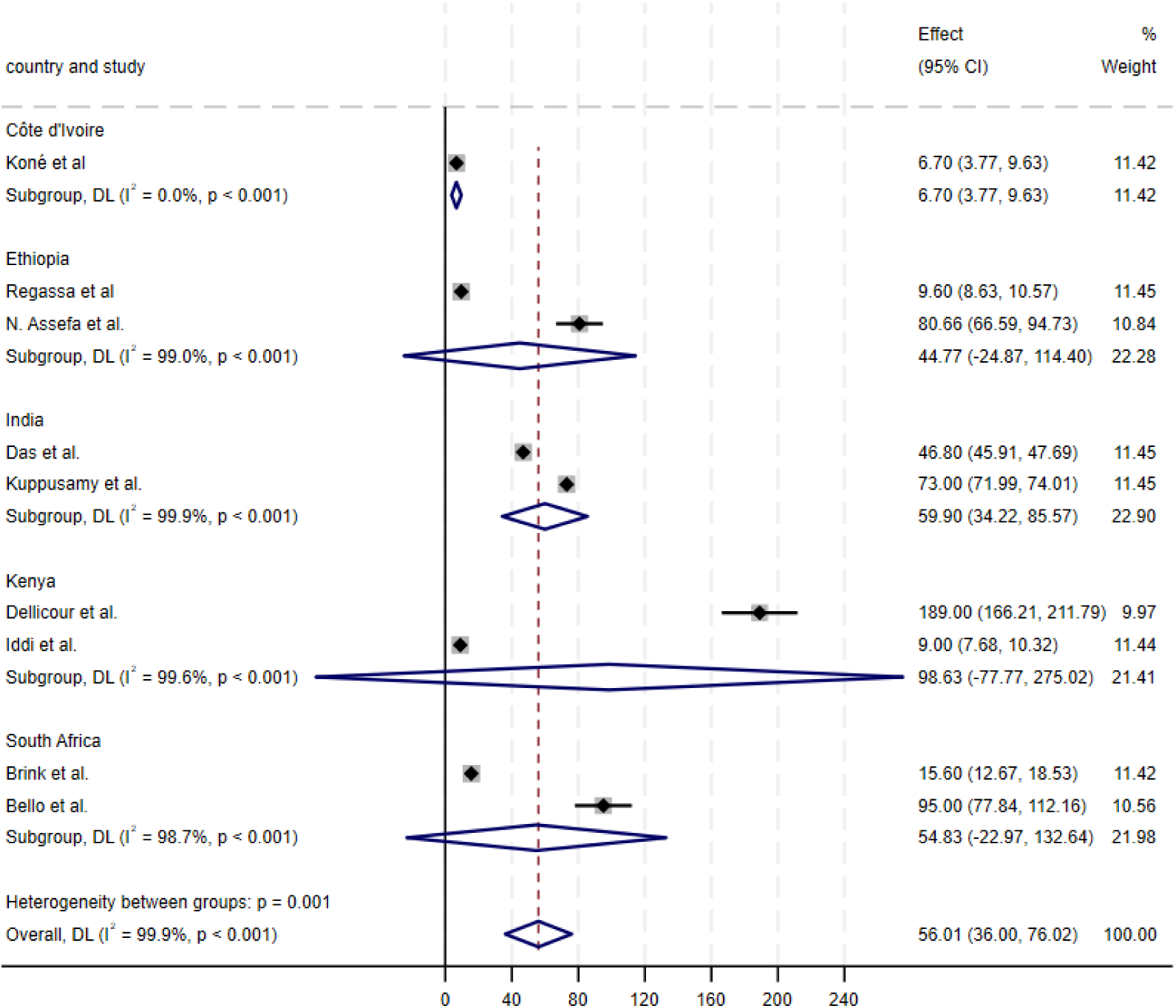
Meta-analysis of miscarriage rates per 1,000 pregnancies. Notes: Figure shows a meta-analysis of estimates of the rate of miscarriage per 1000 pregnancies from prior studies in the five study countries.

**Figure S3:**
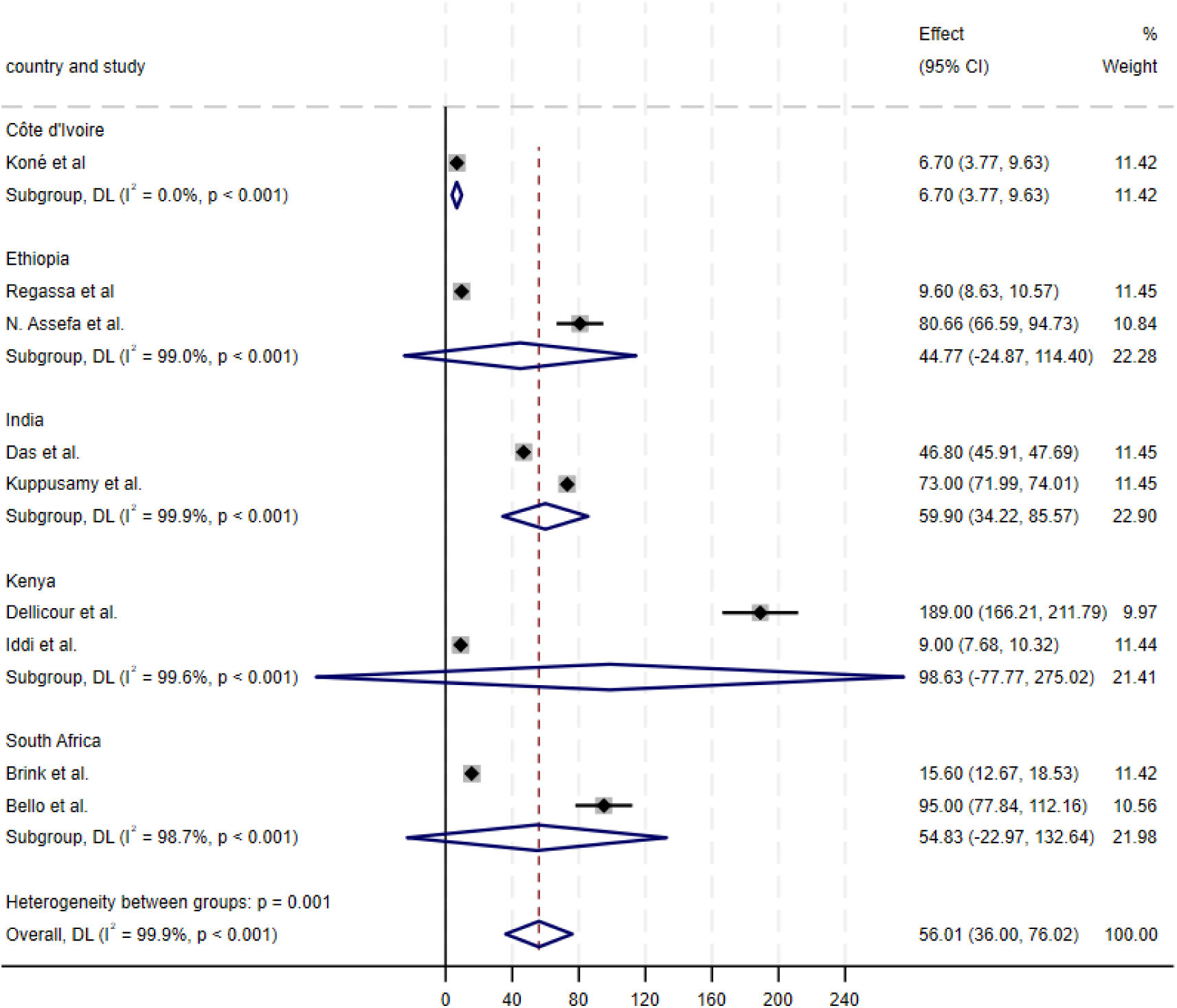
Meta-analysis of stillbirths rates per 1,000 births. Notes: Figure shows a meta-analysis of estimates of the rate of stillbirth per 1000 births from prior studies in the five study countries. When studies report stillbirth per 100 pregnancies, estimates are adjusted for comparability. Several studies report the stillbirth rate per pregnancies rather than per births; these have been adjusted here using data from the published paper so that the rate is reported as the number of stillbirths divided by the number of births, excluding miscarriages from the denominator.

